# A Hybrid Machine Learning Framework for Enhancing the Prediction Power in Large Scale Population Studies: The ATHLOS Project

**DOI:** 10.1101/2021.01.23.21250355

**Authors:** Petros Barmpas, Sotiris Tasoulis, Aristidis G. Vrahatis, Matthew Prina, José Luis Ayuso-Mateos, Jerome Bickenbach, Ivet Bayes, Martin Bobak, Francisco Félix Caballero, Somnath Chatterji, Laia Egea-Cortés, Esther García-Esquinas, Matilde Leonardi, Seppo Koskinen, Ilona Koupil, Andrzej Pająk, Martin Prince, Warren Sanderson, Sergei Scherbov, Abdonas Tamosiunas, Aleksander Galas, Josep Maria Haro, Albert Sanchez-Niubo, Vassilis Plagianakos, Demosthenes Panagiotakos

## Abstract

The ATHLOS cohort is composed of several harmonized datasets of international cohorts related to health and aging. The healthy aging scale has been constructed based on a selection of particular variables from 16 individual studies. In this paper, we consider a selection of additional variables found in ATHLOS and investigate their utilization for predicting the healthy aging. For this purpose motivated by the dataset’s volume and diversity we focus our attention upon the clustering for prediction scheme, where unsupervised learning is utilized to enhance prediction power, showing the predictive utility of exploiting structure in the data by clustering. We show that imposed computation bottlenecks can be surpassed when using appropriate hierarchical clustering within a clustering for ensemble classification scheme while retaining prediction benefits. We propose a complete methodology which is evaluated against baseline methods and the original concept. The results are very encouraging suggesting further developments in this direction along with applications in tasks with similar characteristics. A strait-forward open source implementation is provided for the R project.

## 1. Introduction

Health informatics has received much attention in the past few years since it permits big data collection and analytics and extracts patterns that are free of the strict methodological assumptions of statistical modeling [1, 2]. Recent advances in the biomedical domain generate data at an increasing rate in which approaches under the perspective of health informatics, contribute in the accurate early disease detection, patient care, and community services. These complex data belong to the “Big data” category containing various variable types with different scales or experimental setups, in many cases incomplete [3]. The large data volume on each biomedical research field offers the opportunity to open new avenues for exploring the various biomedical phenomena. Machine learning methods are considered as the first choice for the analysis of this data as they can manage their volume and complexity. In recent years both unsupervised and supervised machine learning methods have been applied to biomedical challenges offering reliable results.

A large category on this perspective is the population studies for aging and health analysis where they offer a plurality of large scale data with high diversity and complexity. Aging and health indicators are an important part of such research as the population aging observed in most developed countries leads to an increasing interest in studying health and aging, since the elderly are nowadays the fastest-growing segment in large regions, such as Europe, Asia and the USA [4, 5, 6, 7, 8]. As such, discovering health-related factors in an attempt to understand how to maintain a healthy life is of crucial importance. Meanwhile, it has long been reported that Sociodemographic factors are significant determinants of various health outcomes such as healthy aging [9, 10], while evidently aging involves interactions between biological and molecular mechanisms with the environment, and as a result, it is a multifactorial phenomenon that everyone experiences differently [11].

The EU-funded ATHLOS (Ageing Trajectories of Health: Longitudinal Opportunities and Synergies (EU HORIZON2020–PHC-635316, http://athlosproject.eu/)) Project produces a large scale dataset in an attempt to achieve a better understanding of ageing. The produced harmonized dataset includes European and international longitudinal studies of aging, in order to identify health trajectories and determinants in aging populations. Under the context of ATHLOS, a metric of health has been created using an Item Response Theory (IRT) approach [12] delivering a common metric of health across several longitudinal studies considered in ATHLOS. Interestingly, there is a plethora of available variables within the harmonized dataset that have not been considered when generating the aforementioned metric of health, encouraging the further exploration of associated factors through the utilization of Pattern Recognition and Machine Learning (ML) approaches. Nevertheless, the imposed data volume and complexity generate challenges for ML related to big data management and analytics.

There is a plethora of recently published studies based on the ATHLOS dataset with promising results in several fields. Such fields include cardiovascular disease evaluation [13, 14, 15, 16, 17], demographic studies about sociodemographic indicators of health status [18] and the impact of socioeconomic status [19, 20, 21], nutrition science studies such as nutrition effects on health [22, 23, 24] and alcohol drinking patterns effects on health [25, 26] and even psychology studies assessing the impact of depression and other psycological disorders related to aging and health [27, 28, 29]. Nevertheless, the ATHLOS data specifications require analysis through Machine Learning methods to uncover the data complexity and better interpreting the characteristics that affect the state of human health. Predicting the health index can be considered one of the greatest challenges of ATHLOS projects in the health informatics domain. Previously, members of the ATHLOS consortium published studies [30, 31] by applying various supervised Machine Learning algorithms on part of ATHLOS data (ATTICA and ELSA study respectively). While these studies have shown remarkable results, a study of the health status prediction in the unified and harmonized ATHLOS data utilizing all additional information has not yet been done.

In this study, we proposed a hybrid machine learning framework which includes the integration of Unsupervised and Supervised Machine Learning Algorithms to enhance prediction performance on large-scale complex data. More precisely, we developed a divisive hierarchical clustering for ensemble learning framework to enhance the prediction power on ATHLOS large-scale data regarding its Health Status score. We focus our attention upon the clustering for prediction scheme, where unsupervised learning is utilized to enhance prediction power, showing the predictive utility of exploiting structure in the data by clustering. We show that imposed computation bottlenecks can be surpassed when using appropriate hierarchical clustering within a clustering for ensemble classification scheme while retaining prediction benefits. We propose a complete methodology which is evaluated against baseline methods and the concept’s basis. The results are very encouraging suggesting further developments in this direction along with applications in tasks with similar characteristics.

## 2. Related Work

In the last decade, several studies have been published regarding the integration of unsupervised and supervised learning strategies, most of which concern the incorporation of clustering models to classification algorithms for the improvement of the prediction performance. Although there has been a remarkable progress in this area, there is a need for more robust and reliable frameworks under this perspective given the ever-increasing data generation in various domains. Clustering can be considered as a pre-processed step in a classification task since in complex data with non-separable classes the direct application of a classifier can be ineffective. In [32] the authors provided evidence that the training step in separated data clusters can enhance the predictability of a given classifier. In their approach the k-means and a hierarchical clustering algorithm were utilized to separate the data while neural networks were applied for the classification process.

The utility of clustering in gaining more information about the data and subsequently reducing errors in various prediction tasks has been previously explored, with promising outcomes in various domains. The clustering outcome can be considered as a dataset’s compressed representation which has the potential to exploit information about the data and its structure, further employed to improve the predictive power. In [33] the authors examine the extent to which analysis of clustered samples can match predictions made by analyzing the entire dataset at once. For this purpose, they compare prediction results using regression analysis on original and clustered data. It turned out that, clustering improved regression prediction accuracy for all examined tasks. Additionally, the authors in [34] also investigated whether clustering can improve prediction accuracy by providing the appropriate explanations. The proposed a process which concerns the coordination of multiple predictors through a unified ensemble scheme. Furthermore, in [35], the authors integrated the semi-supervised fuzzy c-means (SSFCM) algorithm into the support vector machines (SVM) classifier offering promising results regarding the improvement of SVM prediction power. Their hypothesis lies on the fact that unlabeled data include an inner structure which can be efficiently uncovered by data clustering tools, a crucial step to enhance the training phase of a given classifier. Following a similar perspective, the SuperRLSC algorithm utilizes a supervised clustering method to improve classification performance of the Laplacian Regularized Least Squares Classification (LapRLSC) algorithm [36]. Their motivation is based on the intuition that the clustering process contributes to the identification of the actual data structure by constructing graphs which can reflect more refined data structure. A step further is to incorporate ensemble clustering before the classification stage since an ensemble approach can elucidate the data structure in a more realistic manner [37]. The authors applied this framework to identify breast cancer profiles providing reliable results since ensemble clustering algorithms can deal with the biological diversity is extremely important for clinical experts. Other approaches such as the work in [38] utilize the clustering process to reduce the number of instances used by the imputation on incomplete datasets. The unsupervised learning part in this method offered better results not only in the classification accuracy but also in terms of computational execution time. Given that the most population-based studies include a plethora of missing values, this framework has a great potential to export reliable results in cases. Although several hybrid approaches including supervised and unsupervised machine learning techniques have been recently proposed, the rise of Big Data challenges along with the diversity issues on population studies, necessitates further developments in this direction.

## 3. Background Material

### 3.1. Ensemble Learning

Ensemble methods have seen rapid growth in the past decade within the machine learning community [39]. An ensemble is a group of predictors, each of which gives an estimate of a response variable. Ensemble learning is a way to combine these predictions with the goal that the generalization error of the combination is smaller than each of the individual predictors. The success of ensembles lies in the ability to exploit the diversity in the individual predictors. That is, if the individual predictors exhibit different patterns of generalization, then the strengths of each of the predictors can be combined to form a single, more reliable one.

A significant portion of research outcomes in ensemble learning aims towards finding methods that encourage diversity in the predictors. Mainly, there are three reasons for which ensembles perform better than the individual predictors [40]. The first reason is statistical. A learning algorithm can be considered a way to search the hypotheses space to identify the best one in it. The statistical problem is caused due to insufficient data. Thus, the learning algorithm would give a set of different hypotheses with similar accuracy on the training data. With ensembling, the risk of choosing the wrong hypothesis would be averaged out to an extend. The second reason is of computational nature. Often, while looking for the best hypothesis, the algorithm might be stuck in local optima, thus giving the wrong result. By considering multiple such hypotheses, we can obtain a much better approximation to the true function. The third reason is representational. Sometimes the true function might not be any hypothesis in the hypotheses space. With the ensemble method, the representational space might be expanded to give a better approximation of the true function.

Ensemble learning also coincides with the task of clustering since the performance of most clustering techniques is highly data-dependent. Generally, there is no clustering algorithm, or the algorithm with distinct parameter settings, that performs well for every set of data [41]. To overcome the difficulty of identifying a proper alternative, the methodology of cluster ensemble has been continuously developed in the past decade.

### 3.2. Projection Based Hierarchical Divisive Clustering

Hierarchical clustering algorithms construct hierarchies of clusters in a top-down (divisive) or bottom-up (agglomerative) fashion. The former starts from *n* clusters, where *n* stands for the number of data points, each containing a single data point and iteratively merge the clusters to satisfy certain closeness measures. Divisive algorithms follow a reverse approach, starting with a single cluster containing all the data points and iteratively split existing clusters into subsets. Hierarchical clustering algorithms have been shown to result in high-quality partitions. Nonetheless, their high computational requirements usually prevent their usage in big data scenarios. However, more recent advancements in both agglomerative [42, 43] and divisive strategies [44, 45] have exposed their broad applicability and robustness. In particular, it has been shown that, when divisive clustering is combined with integrated dimensionality reduction [46, 47, 48], we can still get methods capable of indexing extensive data collections. In contrast to agglomerative methodologies, such indexes allow fast new sample allocation to clusters.

In more detail, several projection-based hierarchical divisive algorithms try to identify hyper-planes that best separates the clusters. This can be achieved with various strategies, more notably by calculating the probability distribution of the projected space and avoid separating regions with high-density [49, 50, 51]. The latter though, oppose computational challenges in the density calculation of each neighborhood of high density. Motivated by the work of [52], instead of finding the regions with high density, the authors in [46, 48] try to identify regions with low density to create the separating hyper-planes.

The dePDDP [46] algorithm builds upon “principal direction divisive partitioning” [53], which is a divisive hierarchical clustering algorithm defined by the compilation of three criteria, for the cluster splitting, cluster selection, and termination of the algorithm respectively. These algorithms incorporate information from the projections *p*_*i*_: *p*_*i*_ = *u*_1_ (*d*_*i*_ − *b*), *i* = 1, …, *n* onto the first principal component *u*_1_ to produce the two subsequent partitions at each step. In more detail, dePDDP splits the selected partition 𝒫^↕^ by calculating the kernel density estimation 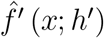 of the projections 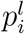 and the corresponding global minimiser *x*^∗^ defined as the best local minimum of the kernel density estimation function. Then constructs 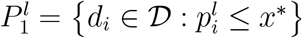 and 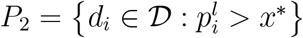. Now, let 𝒫 a partition of the dataset *D* into *k* sets. Let *F* be the set of the density estimates 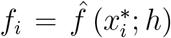 of the minimisers 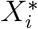 for each *C*_*i*_ ∈ 𝒫. The next set to split is *C*_*j*_, with *j* = arg max_*i*_ {*f*_*i*_ : *f*_*i*_ ∈ ℱ} Finally, the algorithm allows the automatic determination of clusters by terminating the splitting procedure as long as there are no minimiser for any of the clusters *C*_*i*_ ∈ 𝒫.

By using techniques like the fast Gauss transform, linear running time for the kernel density estimation is achieved, especially for the one-dimensional case. To find the minimiser, only the density at n positions needs to be evaluated, in between the projected data points, since those are the only places with valid splitting points. Thus, the total complexity of the algorithm remains O (*k*_max_ (2 + *k*_*SVD*_) (*s*_*nz*_*na*)).

Minimum Density Hyper-planes (MDH) algorithm [48] follows a similar clustering procedure, however, instead of using the First Principal Component for the calculation of the splitting hyper-plane that minimizes the density, follows a projection pursuit formulation of the associated optimization problem to find minimum density hyper-planes. Projection pursuit methods optimise a measure of interest of a linear projection of a data sample, known as the projection index, in this case the minimum value of the projected density. Although this is a theoretically justified approach, it is more computationally intensive mainly due to the optimization procedure as such when either clustering efficiency is not of crucial importance (data indexing) or computation burden limit applicability, the dePDDP approach can be consider as a satisfactory approximation of MDH.

## 4. The Proposed Ensemble Methodology

The concept proposed in [34] showed that an ensemble learning predictor based on different clustering outcomes can improve the prediction accuracy of regression techniques. The performance gains are associated with the change in locality features when training prediction models for individual clusters, rather than the whole dataset. Different clustering outputs 𝒫 are retrieved by providing various *k* values to the *k*-means clustering algorithm increasing the diversity of the outcomes. For *k* = 1 … *L* we retrieve *L* 𝒫_*k*_ individual partitionings. Then for each cluster 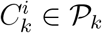 with *i* = 1 … *k* and *k* = 1 … *L*, a model is trained. The final predictions for each data point are calculated by averaging amongst the predicted values retrieved by the models that correspond to the clusters 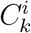 that falls within. Selecting a cutoff *L* for *k* (how many individual partitionings 𝒫_*k*_ should be calculated) is not clear but data dependent heuristics can estimated.

There is a crucial trade-off, however, for this methodological framework with respect to the computational complexity, imposed by the number of predictors that need to be trained. Even though each model is trained upon a subset of the original dataset, we still need to train 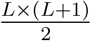 predictors. As a result, the computational complexity expresses exponential behavior. Large scale prediction tasks similar to the one studied here can prohibit the extensive utilization of this concept in particular when combined with computationally demanding predictors such as Neural Networks and Support Vector Machines.

In this work, motivated by recent advantages in projection-based divisive hierarchical algorithms, we proposed an ensemble algorithmic scheme able to surpass the aforementioned computational burden while retaining prediction benefits. The key idea is to generate the *L* partitionings by iteratively expanding a binary tree structure. Divisive clustering algorithms allow us to stop the clustering procedure as long as the predefined number of clusters *k* has been retrieved. Then to retrieve the partitioning for *k* = *k* + 1 we only need to split one of the leaf nodes. In practice, all partitionings *L* can be retrieved by a single execution of the algorithm where *k* is set to the threshold value *L*. By monitoring the order of binary splits we retrieve 𝒫 constituted by the individual partitionings that correspond to the *k* = 1 … *L* values. Arguably, we sacrifice some of the diversity between the individual partitionings 𝒫 _*k*_ since each two consecutive partitionings only differ with respect the portion of the dataset that constitutes the selected for splitting leaf node, but simultaneously benefit greatly by only having to train 2*L* + 1 models. Again, to provide the final prediction for each data point we need to average the predicted values retrieved by the models that correspond to the clusters 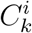. This means that we need to combine information retrieved by the nodes (clusters) appearing along the path each sample followed from the root node (containing the full dataset) the the leaf node that lies within. Note that this divisive structure not only allow us to interpret the ensemble procedure, but it is also strait forward to efficiently assign new observations to the tree structure providing the corresponding predictions for new arriving samples.

### Algorithm 1: Clustering for Ensemble Prediction Framework

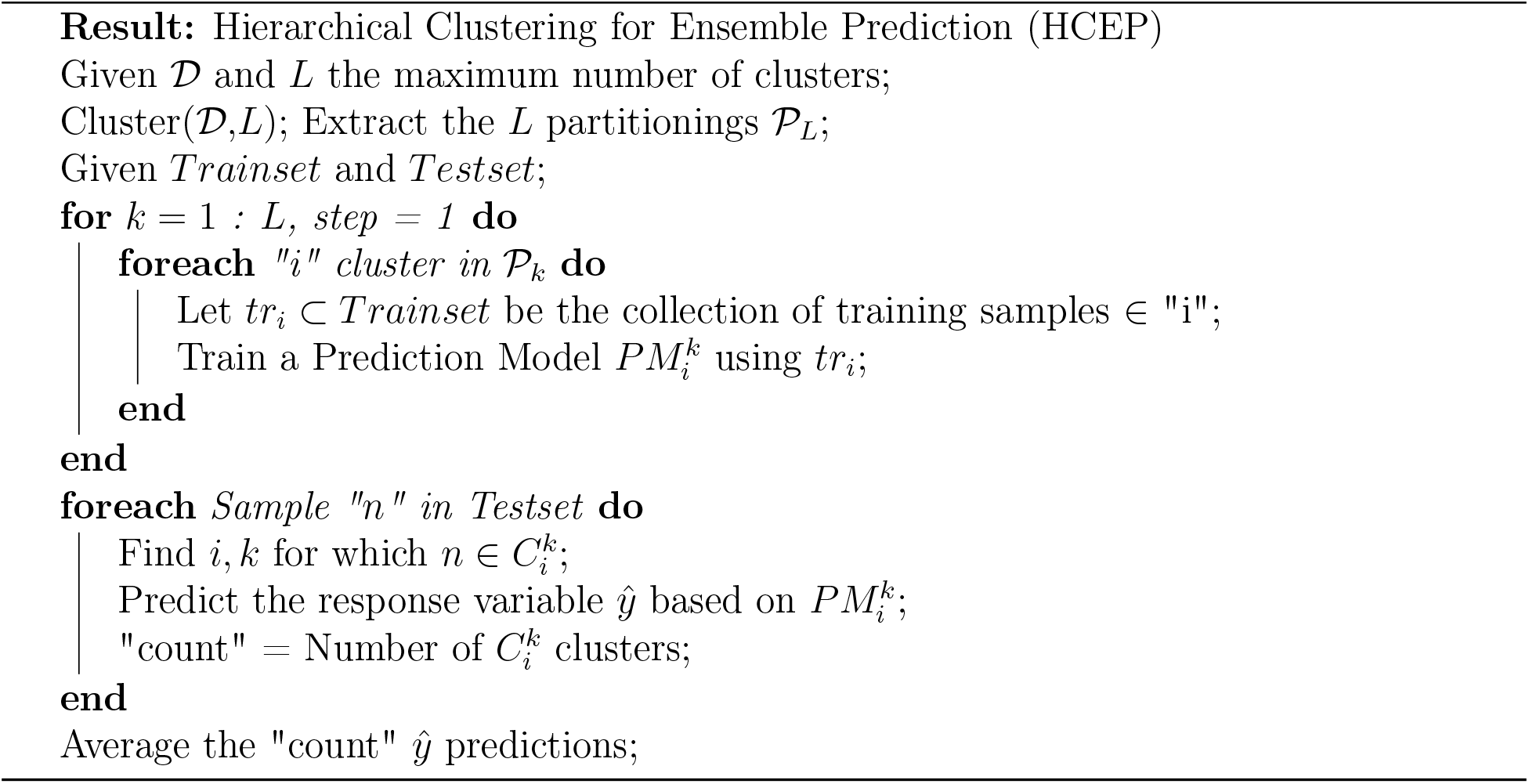

In Algorithm 1 we present the complete proposed algorithmic procedure entitled “Hierarchical Clustering for Ensemble Prediction (acronym: HCEP)”. In summary, the first step is to execute the projection based divisive clustering algorithm of choice and retrieve the complete resulting binary clustering tree. Keep in mind that the response variable is not talking into account for this step, as such, this is an unsupervised procedure.Then for each node of the tree we train the selected prediction algorithm based only on samples belonging to the train set. For every sample belonging to the test set we can now provide final predictions by averaging across the individual predictions of this particular sample retrieved by the corresponding nodes of the tree that lies within. For each new arriving sample we initially pass it through the tree structure until reaching the appropriate leaf node. This is done by projecting the new sample onto the one dimensional vector retrieved for each node of the tree and deciding whether it should be assigned at the right or the left child further on. Then the prediction mechanism is applied as before.

### 4.1. Naive Clustering for Prediction

We are also interested in investigating the effectiveness of clustering in prediction when used as a single pre-processing step [33]. We expect that the characteristics of the ATHLOS dataset employed in this work, such as its large scale and the imposed complexity by the appearance of both continuous and categorical variables, present a unique opportunity to expose the benefits, if any, in training individual models for sub-populations of samples belonging to the same cluster.

In practice, this procedure can be achieved utilizing any clustering algorithm. Here we employ both k-means and projection based divisive clustering as representatives of partitioning and hierarchical clustering respectively, that also allow straightforward allocation of new arriving samples to retrieved clusters. The algorithmic procedure is presented in Algorithm 2. The clustering takes place initially for a given number of clusters which is subject to further investigation, then a prediction model is trained for each cluster utilizing the respective train samples, while for each sample in the train set, the final prediction is provided by the model that corresponds to the cluster it lies within. The new arriving sample are initially allocated to a cluster and then a similar procedure is followed to provide predictions. Notice that, this procedure should be significantly more computationally efficient than the ensemble methodology since we only need to train *L* models. In addition, for particular prediction algorithms with close to exponential complexity with respect to the number of samples, we also expect a significant computational boost against their application on the full dataset 𝒟.

#### Algorithm 2: Clustering for Prediction Framework

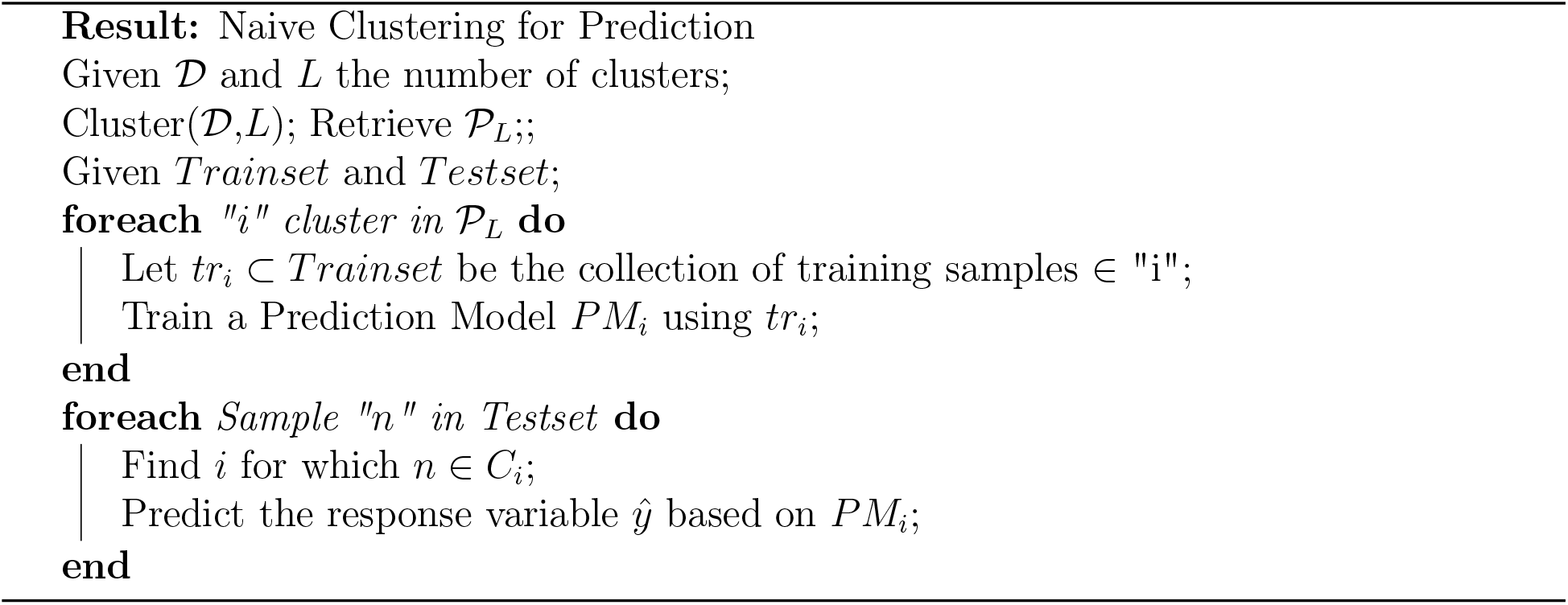

## 5. Data Description and Pre-processing

The ATHLOS harmonized dataset [54] includes European and international longitudinal studies of aging. It contains more than 355,000 individuals who participated in 17 general population longitudinal studies in 38 countries. We specifically used 15 of these studies, which are: 10/66 Dementia Research Group Population-Based Cohort Study [55], the Australian Longitudinal Study of Aging (ALSA) [56], Collaborative Research on Ageing in Europe (COURAGE) [57], ELSA [58], Study on Cardiovascular Health, Nutrition and Frailty in Older Adults in Spain (ENRICA), [59], the Health, Alcohol and Psychosocial factors in Eastern Europe Study (HAPIEE), [60], the Health 2000/2011 Survey [61], HRS [62], JSTAR [63], KLOSA [64], MHAS ([65]), SAGE [66], SHARE, [67], the Irish Longitudinal Study of Ageing (TILDA) [68] and the the Longitudinal Aging Study in India (LASI) [69]. The aforementioned studies consist of 990,000 samples with more than 355,000. The dataset contains 184 variables, two response variables, and 182 independent variables. Response variables are the raw and the scaled Healthstatus scores of each patient. Regarding the independent variables (see supplementary material sheet S1), nine variables were removed including various indexes (sheet S2), 13 variables were removed including obviously depended variables that cannot be taken into account (sheet S3), and six variables were removed including information that cannot be considered within the prediction scheme (sheet S4). Furthermore, the 47 variables (sheet S5), which originally calculate the HS score [12] are excluded. No only, these features create a statistical bias regarding the HS, which is the response variable in our analysis, but also, in this work we aim to uncover new insights for external variables that have previously been considered not significantly relevant. Removing any samples for which the HS metric is not available, the resulting data matrix is constituted by 770,764 samples and 107 variables.

To this end, we have to deal with the critical step of missing value imputation. For this purpose we utilized the Vtreat [70] methodology, a cutting-edge imputation tool with reliable results. Vtreat is characterized by a unique strategy for the dummy variables creation which resulted to the construction of 458 dummy variables in total. Next a significance pruning process step took place where each variable was evaluated based on its correlation with the HealthStatus score (response variable).

## 6. Experimental Analysis

In the first part of our experimental analysis, we compare the proposed ensemble scheme based on Projection Based Hierarchical Clustering (HCEP) against the original one, based on *k*-means partitioning clustering. We also examine if there are any benefits when compared against the naive clustering for prediction scheme presented in Section 4.1, utilizing both aforementioned clustering approaches. For this set of experiments the divisive algorithm of choice is dePDDP, while the maximum number of clusters *L* is set to 40, a value greater than the average optimal number of clusters retrieved by dePDDP, to effectively examine the methodology’s behaviour. For every run of *k*-means and dePDDP the number of clusters *k* is given as input. *k*-means is allowed to choose the most appropriate convergence amongst 10 random initializations [71, 72], while for dePDDP the “bandwidth multiplier parameter” is set to 0.05, a relative small value to guaranty enough binary splits that will lead to the required number of leafs (clusters). Finally, to avoid highly unbalanced tree structures we set a threshold, so that clusters with less than *N/k* points are not allowed to be split [73], where *N* is the total number of points in the dataset. All methodologies are implemented for “R-project”, while specifically for dePDDP we utilize a native efficient implementation and for *k*-means we employed the implementation provided by the “biganalytics” package called BigKmeans [74], which benefit from the lack of memory overhead by not duplicating the data. This choice for the employed clustering algorithms is based not only on their satisfying performance but also on their simplicity and the structural ability to create an index that can be used to allocate future observations. For dePDDP each new instance pushed into the tree until it reached the respective leaf node. For the Kmeans algorithm, we allocate every instance of the testing set to the closest cluster by calculating the minimum distance to the cluster centroids.

For the prediction task, we employ the traditional Linear Regression (LR) and Random Forests (RF). Again, the default parameter values are those provided by the corresponding implementations found in [75]. For RF, we used 50 trees to guaranty its low computational complexity, due to restrictions imposed by hardware capabilities, and the *M*_*try*_ variable was defined as *p/*3, where *p* are the number of variables. The regression performance is evaluated with respect to the Root Mean Square Error and the R-squared (RSQ). The mean squared error (MSE) is a measure of an estimator’s quality, with values closer to zero indicating better performance. The MSE is the second moment of the error. Thus, it incorporates both the variance of the estimator (how widely spread the estimates are from one data sample to another) and its bias (how far off the average estimated value is from the truth). MSE has the same units of measurement as the square of the quantity being estimated. In an analogy to standard deviation, taking the square root of MSE yields the root-mean-square error RMSE [76]. R-squared (R2) is a statistical measure that represents the proportion of the variance for a dependent variable that’s explained by an independent variable or variables in a regression model. 

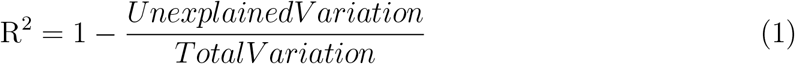

R-squared explains to what extent the variance of one variable explains the variance of the second variable. So, if the R2 of a model is 0.50, then approximately half of the observed variation can be explained by the model’s inputs.

The results with respect to the RMSE metric regarding the prediction of Health Status are reported in Figure 1. To achieve robust validation of the results while maintaining reasonable execution times, we utilize a bootstrapping technique by randomly sampling 50000 samples for training and 1000 samples for testing with replacement [77]. The procedure is repeated 10 times with different subsets for training and testing respectively. Then, we estimate each model’s performance by computing the the average score and the corresponding standard deviation. These are reported using line plots for mean values and shaded ares for standard deviation respectively. The top row of figures corresponds to the naive methodology while the bottom row corresponds to the ensemble approaches respectively. For both cases we report the performance of the catholic models indicated by the straight purple shaded area, parallel to axes X. Orange and Green shaded areas indicate the performance of kmeans and dePDDP algorithms respectively, when combined with either Random Forests (left column) or Linear regression (right column). Notice that performance is reported with respect to the number of clusters (X axes). For the Naive methodology each number of clusters *L* correspond to the RMSE value retrieved for this particular value of *L*, while for the ensemble models for each *L* value we observe the RMSE resulting by aggregating predictions for *k* ∈1 · *L*.

**Figure 1:**
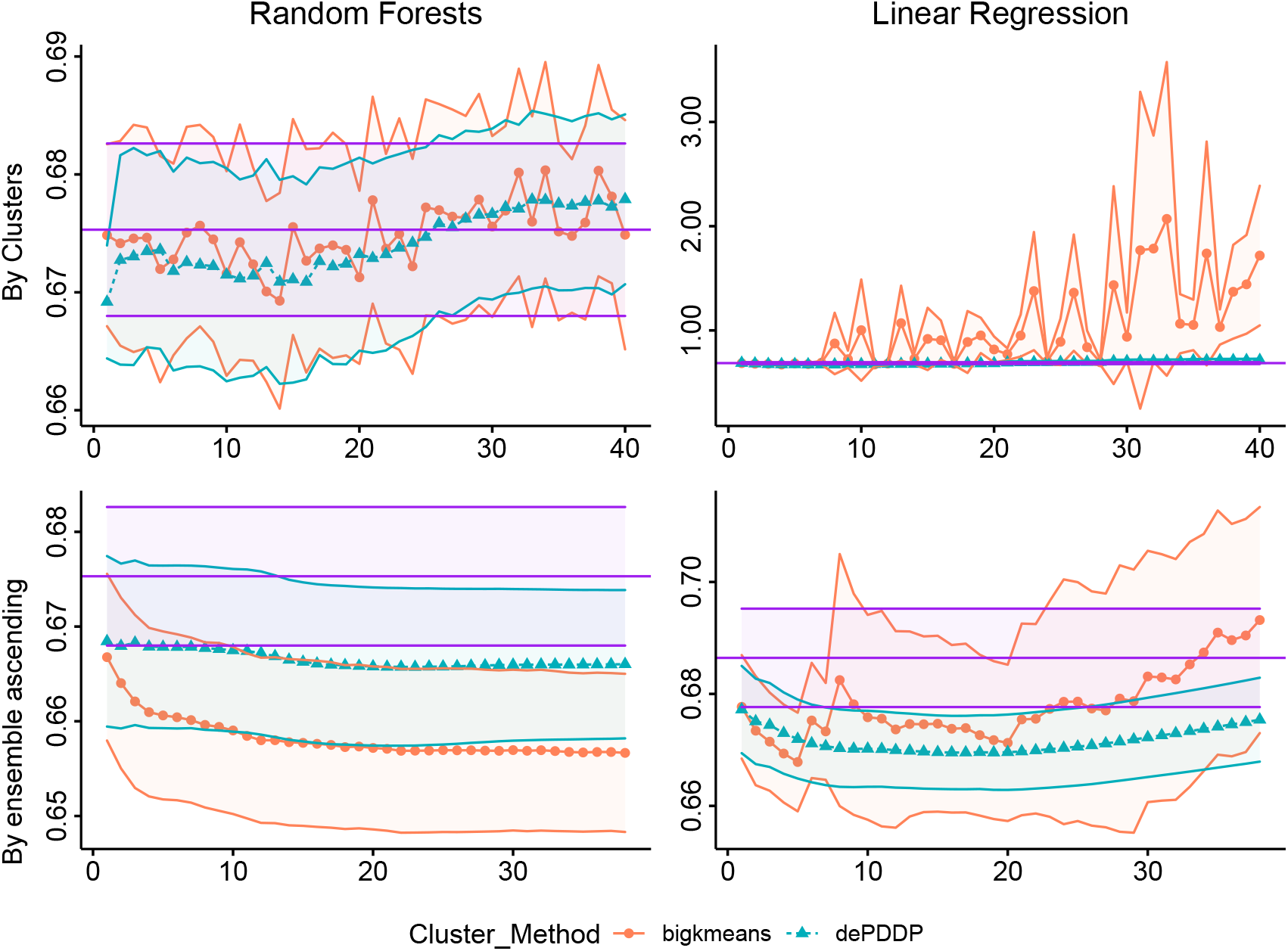
RMSE metric results with respect to the Health Status response variable’s prediction for Random Forests (first column) and Linear Regression (second column) as base regression models. Circular points with continuous red lines represent the results (vertical axes) when bigKmeans algorithm is utilized while, triangular points with blue dashed lines represent the results for dePDDP respectively. Each row of plots depicts the clustering for prediction different strategies. Naive methodology (first row), and Ensemble on ascending range of clusters (second row). The horizontal purple shaded area represents the corresponding values for the catholic models (training a single predictor in the entire dataset). Mean values are reported according to the utilized bootstrapping, while colored ribbons present the standard deviation between the experiments.

In Figure 1 we observe a performance boost compared to the catholic regression models that is more evident and robust for the ensemble methodologies (Algorithm 1). For up to 20 clusters the naive models also appear to improve prediction performance, at least when utilizing RF, but when *k*-means is selected for clustering there is no consistency. For the ensemble models best performance is achieved by *k*-means when combined with RF, while the opposite holds for linear regression. Finally, utilizing dePDDP result in a monotonic behaviour regarding prediction performance in both cases in contrast to *k*-means. Similarly to the naive models, we observe a behaviour indicating an empirical threshold regarding the number of clusters parameter. This is most likely due to over-fitting since for a high enough number of retrieved clusters, we expect to end up with clusters characterized by low sample size compared to the number of variables.

Having concluded that the HCEP framework is able to enhance prediction performance compared to the catholic models and the naive approach, we are also interesting to examine the computational burden. Figure 2 is devoted to the computational time comparisons. As expected, the naive approach can reduce computational at least for complex method such as RF that are greatly affected by samples size. More importantly, we observe the computational complexity comparison between the ensemble approaches, justifying the utilization of the proposed method. It is evident that consistent prediction power benefits can be achieved with minimal computational overhead. Notice here, that the aforementioned computational times for RF have been achieved by implementing a parallel execution strategy accommodated by the “foreach” package [78]. Experiments took place on a PC with Intel i9 processor and 32 GB of RAM running the Ubuntu Linux operating system.

**Figure 2:**
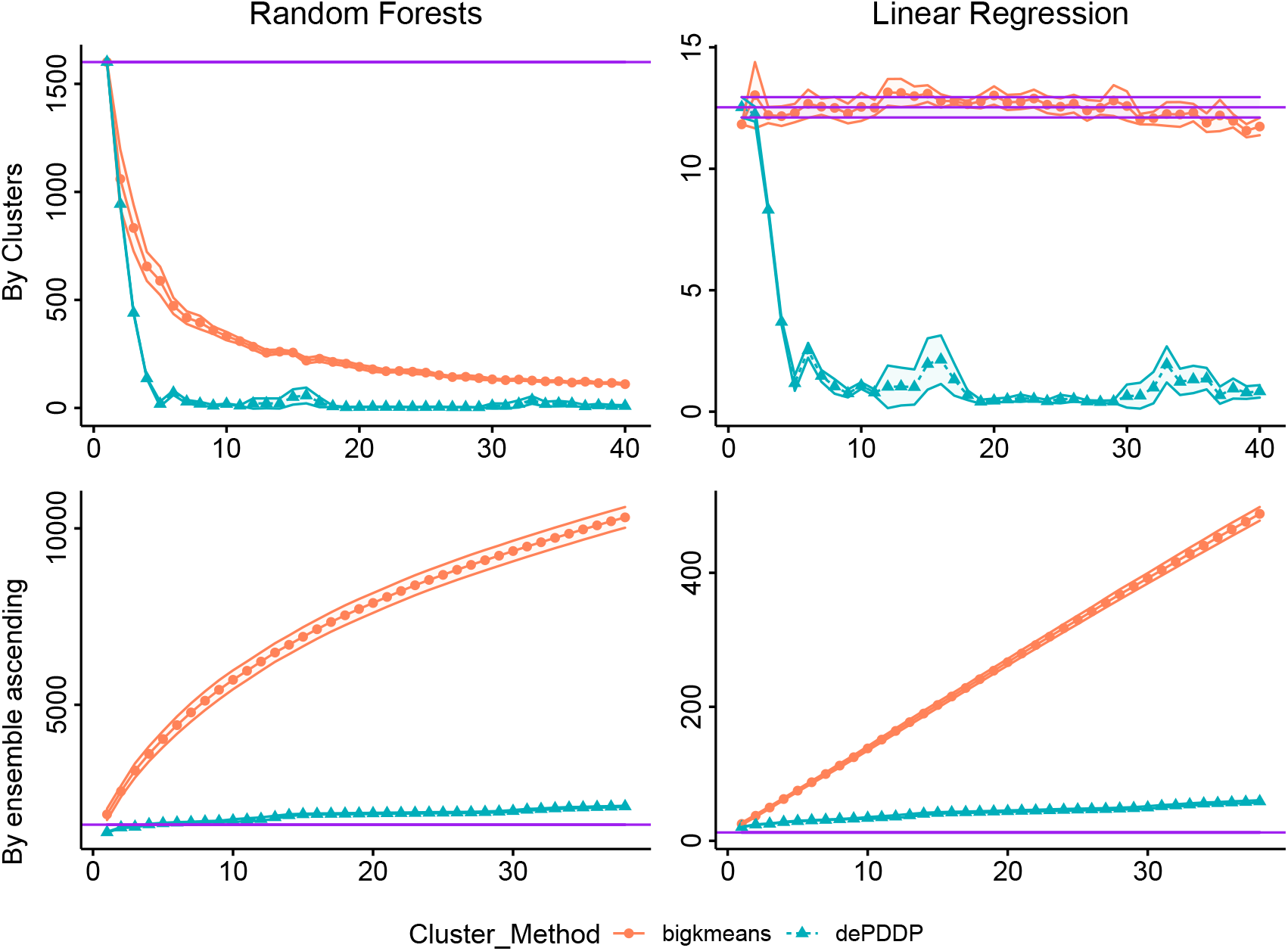
Computational cost in seconds utilizing Random Forest (first column) and Linear Regression (second column) prediction models respectively. Circular points with continuous red lines represent the results (vertical axes) when bigKmeans algorithm is utilized while, triangular points with blue dashed lines represent the results for dePDDP respectively. Each row of plots depicts the clustering for prediction different strategies. Naive methodology (first row), and Ensemble on ascending range of clusters (second row). The horizontal purple shaded area represents the corresponding values for the catholic models (training a single predictor in the entire dataset). Mean values are reported according to the utilized bootstrapping, while colored ribbons present the standard deviation between the experiments.

### 6.1. Extended Comparisons

In what follows, we evaluate the performance of the proposed HCEP methodology comparing it with additional well-established and state-of-the-art regression models in predicting Health Status using the same bootstrapping technique. In detail, six regression models have been applied, namely, the Linear Regression (LR) model, the Random Forests (RF) regression, the k nearest neighbors (kNN) regression, the XGboost [79], and two Deep Neural Network architectures (*DNN*_1_ and *DNN*_2_).

Briefly, in in kNN regression, the average of the HS values of the five Nearest Neighbors of a given test point was calculated. The RF regression performs the RF process by calculating the average of all trees’ output in the final prediction for each test sample. We applied 100 trees and the and the *M*_*try*_ variable was defined as 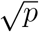, where *p* are the number of variables. Extreme Gradient Boosting (XGBoost) is a cutting-edge classifier based on an ensemble of classification and regression trees [79]. Given the output of a tree *f* (*x*) = *wq*(*x*_*i*_) where *x* is the input vector, and *wq* is the score of the corresponding leaf *q*, the output of an ensemble of K trees will be: 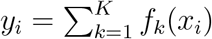.

The first DNN (*DNN*_1_) is constructed with two hidden layers of 100 neurons and one output layer of one neuron, and the second (*DNN*_2_) with one hidden layer with 100 neurons. The ReLU activation function is utilized in hidden layers to control the gradient vanishing problem. The Backpropagation (BP) training algorithm is applied with the learning rate defined as 0.001. We selected these two DNN architectures to deal with both the under- and over-fitting challenges of ATHLOS dataset.

The results are summarized in Table 1. For both ensemble methods we chose to present the values when the maximum number of clusters is set to *L* = 30, which is the average estimated value provided by dePDDP algorithm when utilized for cluster number determination with its default parameters. Notice that, computational limitations do not allow the extensive use of traditional approaches for this purpose[80, 81], while minor variations to this number to not alter the comparison outcome. As shown, the ensemble methodologies outperform any other confirming the prediction enhancement assumption. More precisely, the *k*-means based method combined with RF achieve the best score with respect to both metrics. However, the proposed HCEP performs better when LR is utilized for prediction. In general, HCEP comes second to the original *k*-means based scheme, something to be expected due to the loss of diversity between clusterings as previously discussed in Section 4, however the added value of HCEP arises when considering the minimal computational overhead.

**Table 1:**
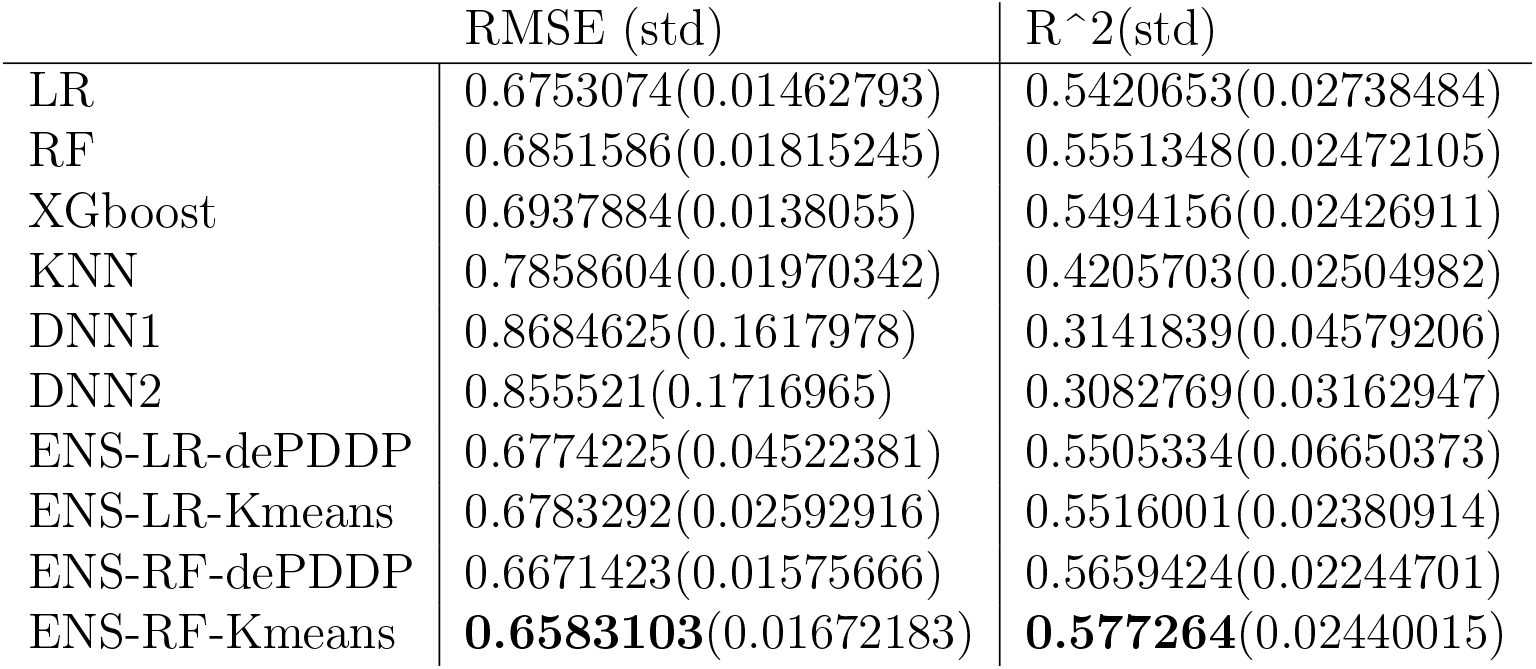
Table presenting the mean RMSE and *R*^2^ for different regression models. The models presented are: Linear Regression (LR), Random Forrests (RF), XGboost, Deep Neural Network with 1 (DNN1) and 2 (DNN2) hidden layers, Hierarchical Ensemble method (HCEP) using Linear Regression or RF based on dePDDP (ENS-LR-dePDDP and ENS-RF-dPDDP respectively) and ensemble method using Linear Regression or RF based on *k*-means (ENS-LR-Kmeans and ENS-RF-Kmeans respectively). In parentheses are the Standard Deviation of the metrics across their 10 individual executions)

| | RMSE (std) | $R^2$(std) |
| --- | --- | --- |
| LR | 0.6753074(0.01462793) | 0.5420653(0.02738484) |
| RF | 0.6851586(0.01815245) | 0.5551348(0.02472105) |
| XGboost | 0.6937884(0.0138055) | 0.5494156(0.02426911) |
| KNN | 0.7858604(0.01970342) | 0.4205703(0.02504982) |
| DNN1 | 0.8684625(0.1617978) | 0.3141839(0.04579206) |
| DNN2 | 0.855521(0.1716965) | 0.3082769(0.03162947) |
| ENS-LR-dePDDP | 0.6774225(0.04522381) | 0.5505334(0.06650373) |
| ENS-LR-Kmeans | 0.6783292(0.02592916) | 0.5516001(0.02380914) |
| ENS-RF-dePDDP | 0.6671423(0.01575666) | 0.5659424(0.02244701) |
| ENS-RF-Kmeans | **0.6583103**(0.01672183) | **0.577264**(0.02440015) |

### 6.2. Tree Visualization and Variable Importance

To visually investigate the clusterability of the dataset at hand through projection based hierarchical clustering we utilized the implementation provided by the R package PPCI [82] (see Figure 3). For this experiment, we utilized HCEP where the maximum number of cluster is conveniently set to *L* = 4. Through the iterative 2d visualization for each node of the tree we visually identify clear patters indicating visually separable clusters. Apparently, the algorithms performs well in identifying clusters, confirming the prediction performance boost we observed previously even for the naive clustering for prediction approach. Note that, sample colouring across the tree structure is following the categorization of the 4 clusters retrieved at the leaf nodes. Finally, we may recall the HCEP procedure using this example. For every train sample that falls in cluster 5 (see Figure 3) the predicted Health Status score is retrieved by averaging the corresponding predictions of the models fitted for clusters 1-3-5.

**Figure 3:**
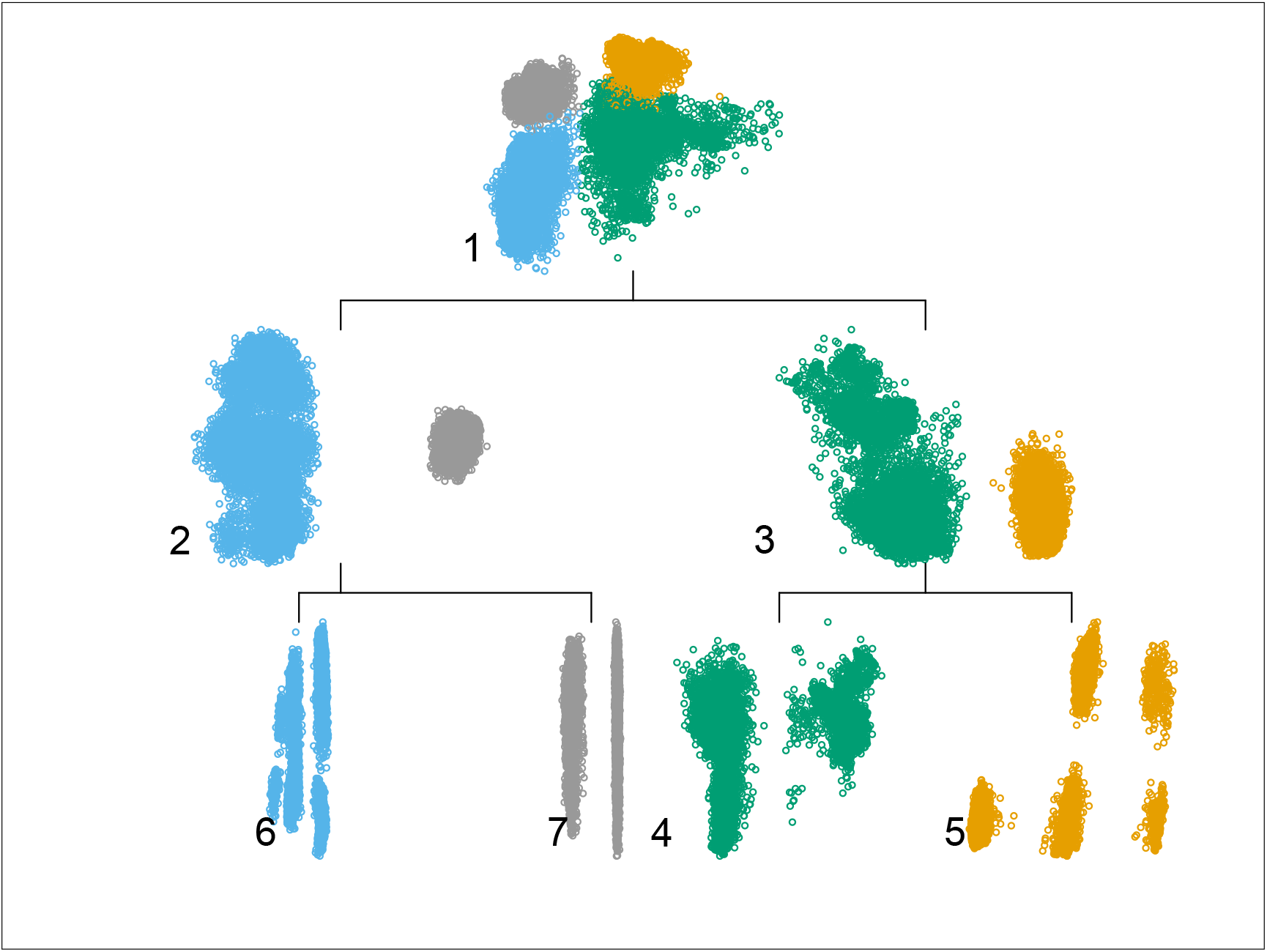
Tree structure example of the hierarchical MDH experiment. In this instance, each step of the algorithm is presented in a top-down order with every level indicating the corresponding cut for a total of 4 clusters. The data points are colored according to the final clustering, and each subset is indicated with numbering from 1 (original dataset) to 6 and 7 (last split producing two of the final clusters).

The straightforward interpretability of the HCEP approach motivated us to further investigate the potential of utilizing it in describing an innovating variable importance analysis. Notice here, that this is an uncommon task for most ensemble prediction approaches or even impossible in many cases. For this purpose we utilized the Percentage Increase in MSE (PiMSE) [75] metric through the Random Forests model for very node of the tree. Then for every path from the root node to each one of the leaf nodes we investigate the PiMSE metric of the nodes within the path since, every point in the test set will be eventually predicted based on one of these paths. For the example at hand (Figure 3) we consider the 10 most important variables, calculated by averaging PiMSE across all aforementioned paths. We illustrate how these differentiate for each one of the four paths 1-3-4, 1-3-4, 1-2-7 and 1-2-6 according to the changes in PiMSE from the root to the lead nodes in Figure 4. In more detail, each subplot depicts one of the four different paths. The PiMSE score is presented in the vertical axes, with the horizontal axes indicating the corresponding node in each path. Larger values in a variable indicate a greater PiMSE score, thus expressing a more significant influence of that variable in that particular node. More specifically, the most important variables depicted here were the “srh” (Respondent’s self-rated/self-reported health, with “*catP* “,”*catN*” etc. implying their transformation variables after the statistical prepossessing), the “h-joint-disorders” (History of arthritis, rheumatism or osteoarthritis), “depression” (Current depressive status) and “age” (Age at time of measure). One example observation we can make through this visualization is that for 2 paths “age” significance drops as tree depth is increasing in contrast to the other two paths for which grows, leading to conclusions such as identifying sub-populations for which a particular variable is relevant in predicting the response variable.

**Figure 4:**
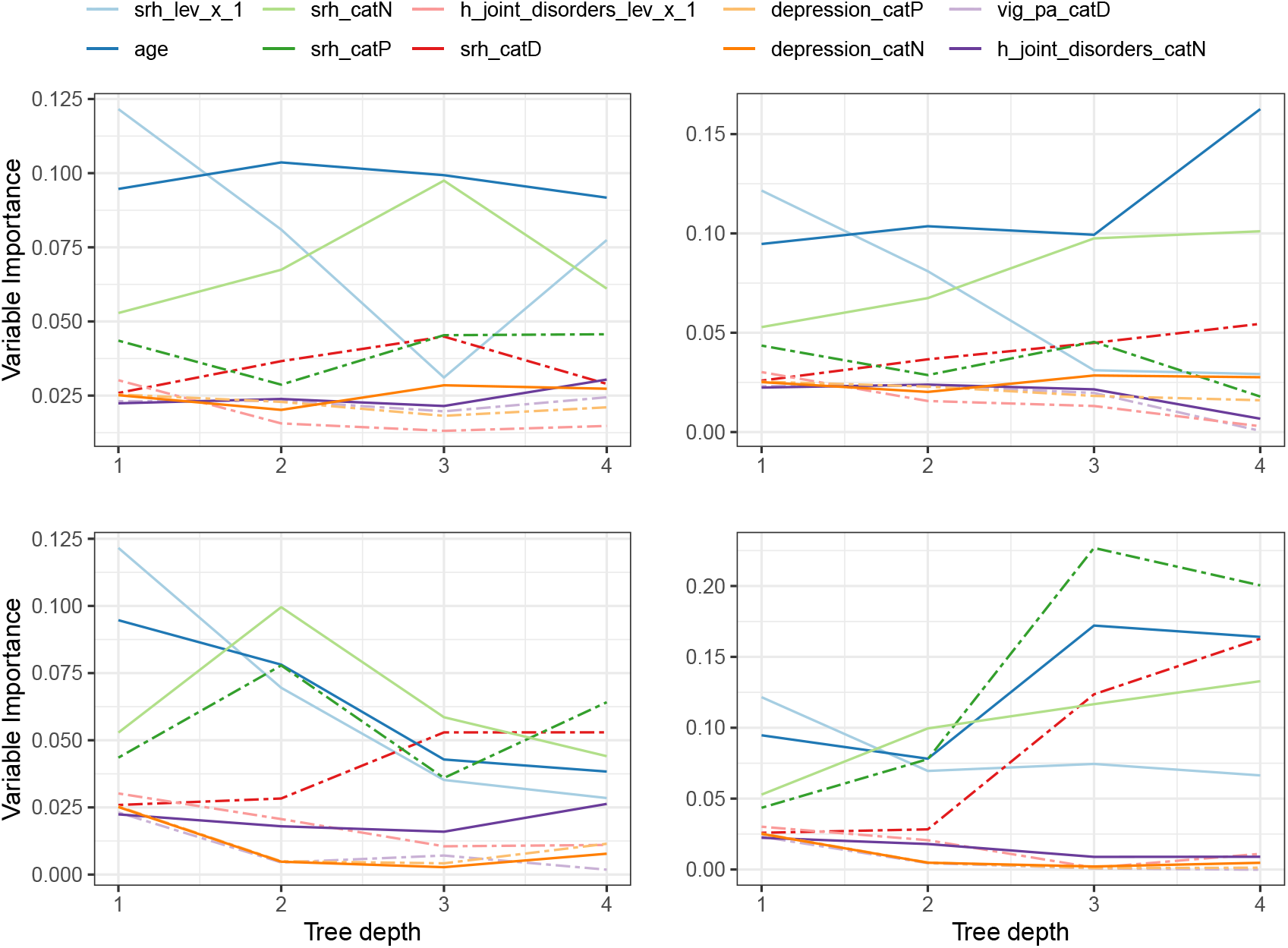
Variable importance propagation of the prediction model’s ten most influencing variables across each node in the different paths of the tree structure.

## 7. Concluding Remarks

Population studies for aging and health analysis offer a plurality of large scale data with high diversity and complexity. Aging and health indicators are an important part of such research, while predicting the health status index can be considered one of the greatest challenges. Motivated by the ATHLOS dataset’s volume and diversity we focus our attention upon the clustering for prediction scheme, where unsupervised learning is utilized to enhance prediction power. We show that imposed computation bottlenecks can be surpassed when using appropriate hierarchical clustering within a clustering for ensemble classification scheme while retaining prediction benefits. In addition, we investigated in depth the interpretability of the proposed architecture exposing additional advantages such as a variable importance analysis. The proposed methodology is evaluated against several regression methods and the original concept with very encouraging results, suggesting further developments in this direction with particular interest in applications with similar characteristics. Thus a strait-forward open source implementation is provided for the R project. The direct expansion of the proposed methodology in classification could suggest a promising future direction, while the utilization of random space transformations to increase diversity of ensemble schemes [83, 84] seems also tempting.

## Data Availability

The data were obtained from the official partners of the ATHLOS project.

http://athlosproject.eu/

## Acknowledgements

This work is supported by the ATHLOS (Aging Trajectories of Health: Longitudinal Opportunities and Synergies) project, funded by the European Union’s Horizon 2020 Research and Innovation Program under grant agreement number 635316.

## Notes

### Competing Interest Statement

The authors have declared no competing interest.

### Author Declarations

Does not apply in our work

### Summary of Updates

Author affiliations updated; Section 1 revised;

